# Predictors and long-term outcomes of oropharyngeal dysphagia in Persons with Multiple Sclerosis: A systematic review protocol

**DOI:** 10.1101/2023.06.15.23291444

**Authors:** Zahra Sadeghi, Mohamadreza Afshar, Asefeh Memarian, Heather L. Flowers

## Abstract

**Background:** Oropharyngeal dysphagia (OPD) can be functionally debilitating in persons with multiple sclerosis (pwMS). OPD induces alterations in safety and efficacy of food and/or liquid ingestion and may incur negative sequalae such as aspiration pneumonia or malnutrition/dehydration. Early detection and timely management of OPD in pwMS could prevent such complications and reduce mortality rates. Identifying predictors of OPD relative to its onset or repeat manifestation will enable the development of care pathways that target early assessment and sustained management. The aims of this systematic review are to compile, evaluate, and summarise the existing literature reporting potential predictors and associated long-term outcomes (e.g., aspiration pneumonia, malnutrition, dehydration, and/or death) of OPD in pwMS.

**Methods:** We will undertake a systematic review to identify studies that describe patterns and complications of OPD in pwMS. Variables of interest include predictors of OPD along with long-term outcomes. We will search MEDLINE, EMBASE, CINAHL, AMED, the Cochrane Library, Web of Science, and Scopus. We will consider studies for inclusion if they involve at least 30 adult participants with MS and report risk factors of OPD and/or its long-term outcomes. Studies will be excluded if they refer to esophageal or oropharyngeal dysphagia induced by causes other than multiple sclerosis. Study selection and data extraction will be performed by two independent assessors for abstract and full article review. We will present study characteristics in tables and document research findings for dysphagia-related risk factors or its complications via a narrative format or meta-analysis if warranted (e.g., mean difference and/or risk ratios measurements). All included studies will undergo risk of bias assessment conducted independently by two authors with consensus on quality ratings.

**Conclusion:** There is a lacune with respect to systematic reviews involving predictors and long-term outcomes of dysphagia in in pwMS to date. Our systematic review will provide the means to develop accurate and efficient management protocols for careful monitoring and evaluation by dysphagia experts. The results of this systematic review will be published in a peer-reviewed journal.

**Systematic review registration:** PROSPERO CRD42022340625

## Introduction

Oropharyngeal dysphagia (OPD) is common in multiple sclerosis (MS) [1] due to injury to the corticobulbar tracts, potentially involving the brainstem, the cerebellum [2,3] and the cortex [4]. It may incur severe and multifaceted poor outcomes, such as aspiration pneumonia, malnutrition/dehydration [3–5], increased psychosocial morbidities [6,7] and even death during periods of medical instability [8]. Identifying predictors for OPD in pwMS will provide the means to develop accurate and efficient management protocols for careful monitoring and evaluation by dysphagia experts. By extension, sustained management will permit timely and sustained care to mitigate potential serious complications.

In two recent systematic reviews, the authors provided an estimate of the pooled frequency of dysphagia in pwMS based on a range of evaluation methods, whether screening, clinical, or instrumental examination [1,9]. Guan et al. [1] reported a pooled frequency estimate of 36% based on subjective screenings or cursory evaluations (such as the Dysphagia in Multiple Sclerosis Questionnaire, the water swallowing test, and various dysphagia check lists from individual clinical swallowing centers) and 81% based on objective measurements (such as videofluoroscopy or fiberoptic nasoendoscopy). More specifically, the frequency of dysphagia was 46% in pwMS when Expanded Disability Status Scale (EDSS) scores were stratified as 4.5 or higher, and 40% for those below 3.0. Similarly, patients with longer disease duration (over 10 years) were more likely to have dysphagia symptoms compared with shorter disease duration.

Various individual studies have also shown a higher frequency of dysphagia with greater disability [10– 14] and/or disease duration [14,15]. Nevertheless, a few studies have reported that pwMS with low EDSS scores still had dysphagia [14,16,17]. To illustrate, Abraham et al. [17] reported that 43% of pwMS had dysphagia whereby 17% of them had low levels of disability (EDSS score lower than 2.5). In contrast, Aghaz et al. [9] estimated the pooled frequency of dysphagia as 37.21% based on subjective evaluations or cursory checklists versus 47% for objective instrumental evaluations respectively. They further studied the presence of dysphagia according to EDSS-based disease severity, duration of disease, and MS stage but failed to demonstrate associations for any of the three factors.

Taken together, reported frequencies of dysphagia hover around one-third of pwMS at a given point in time [1,9]. Varied frequencies might relate to evaluation methods, whether via screening, clinical assessment, or instrumental evaluation. The most common patient-report tool used to identify dysphagia in pwMS is the Dysphagia in Multiple Sclerosis Questionnaire (DYMUS) [1,9], involving 10 items with very good reliability and internal consistency [18]. Frequencies of reported dysphagia based on questionnaires are lower than those based on standardized tools or instrumental evaluations [1]. In general, instrumental assessment remains the gold standard for dysphagia and aspiration detection, whether by videofluoroscopy or fiberoptic nasoendoscopy rather than various types of screening tools or patient reported questionnaires. Some pwMS may underestimate their dysphagia severity due to altered sensory appreciation of symptoms, despite instrumental evidence to the contrary.

In addition to our poor understanding of the frequency of dysphagia in pwMS, gaps exist regarding patterns of associations between disease severity, duration, or stage. Notwithstanding, certain predictive factors may well routinely accompany the expression of dysphagia in pwMS. Elucidating such information would require a comprehensive profile of patient groups with known disease severity, duration, and stage, as well as MS type, neuroanatomical impacts and concomitant deficits or disorders. For example, dysphagia may be precipitated by co-existing psychological or cognitive impairments [10,17,19]. Therefore, continual monitoring for risk of dysphagia in pwMS who also experience negative mental health symptoms or cognitive disorders [4,19] is warranted. Furthermore, dysarthria may be a good and readily identifiable clinical indicator for the presence of dysphagia in persons with neuromuscular diseases [20]. A systematic appraisal of the literature is required to identify the best available evidence for risk factors of dysphagia along with ensuing long-term sequelae in pwMS.

A systematic review constitutes the highest level of research evidence, especially if there is a quality evaluation and meta-analysis. Therefore, establishing the predictors of dysphagia in pwMS, especially when identified with gold standard evaluation methods (such as instrumental assessment), could facilitate the development of new tools for screening or assessing dysphagia and inform practice guidelines. In addition, a close consideration of associated outcomes over the long term (e.g., pneumonia, poor social participation, death) could contribute to our understanding of prognostic indicators for particular patient groups. Consequently, our purpose is to search the existing literature to systematically identify the predictors and associated outcomes of oropharyngeal dysphagia over the long-term in persons with pwMS.

## Methods

The protocol of this systematic review has been registered in PROSPERO (registration number: CRD42022340625). We have applied PRISMA-P guidelines to develop this review protocol further. It serves to direct our search strategy of databases and the grey literature as well as our data extraction and compilation methods. We will document our article selection results using the PRISMA flow diagram to delineate reasons for abstract and article exclusion until the final set of articles is identified. We are submitting the protocol prior to undertaking the full search or any subsequent processes such as abstract screening and full article evaluation.

### Operational Definitions

Dysphagia is defined as body and structure impairment [21] in swallowing evidenced by expert clinical or instrumental assessment of function from the anterior aspect of the lips to inferior aspect of the upper esophageal sphincter. Diagnosis of multiple sclerosis is based on accepted criteria for both definite and probable MS, according to a classification scheme that involves expert clinical and objective evaluations (such as neuroimaging) [22].

### Data sources

We will conduct an electronic search in the following databases for abstracts in languages that the co-authors can read (English, French, German, Persian, Portuguese, Spanish, and Turkish). No publication date or study design restrictions will be imposed. Relevant databases will include MEDLINE, EMBASE, CINAHL, AMED, the Cochrane Library (CENTRAL), Web of Science, and Scopus. The MeSH and search terms used in the search strategy were developed a priori (Table 1). A research librarian will consult to enable valid adaptations of the same terms into the other databases. Our Medline search was conducted in OVID, revealing 189 citations (April 2023). We will also search international grey literature sources (e.g., OpenGrey and Dissertation Abstracts) and review the bibliographies and citations for all included articles in a reiterative manner until no further possible references are identified.

**Table 1.**
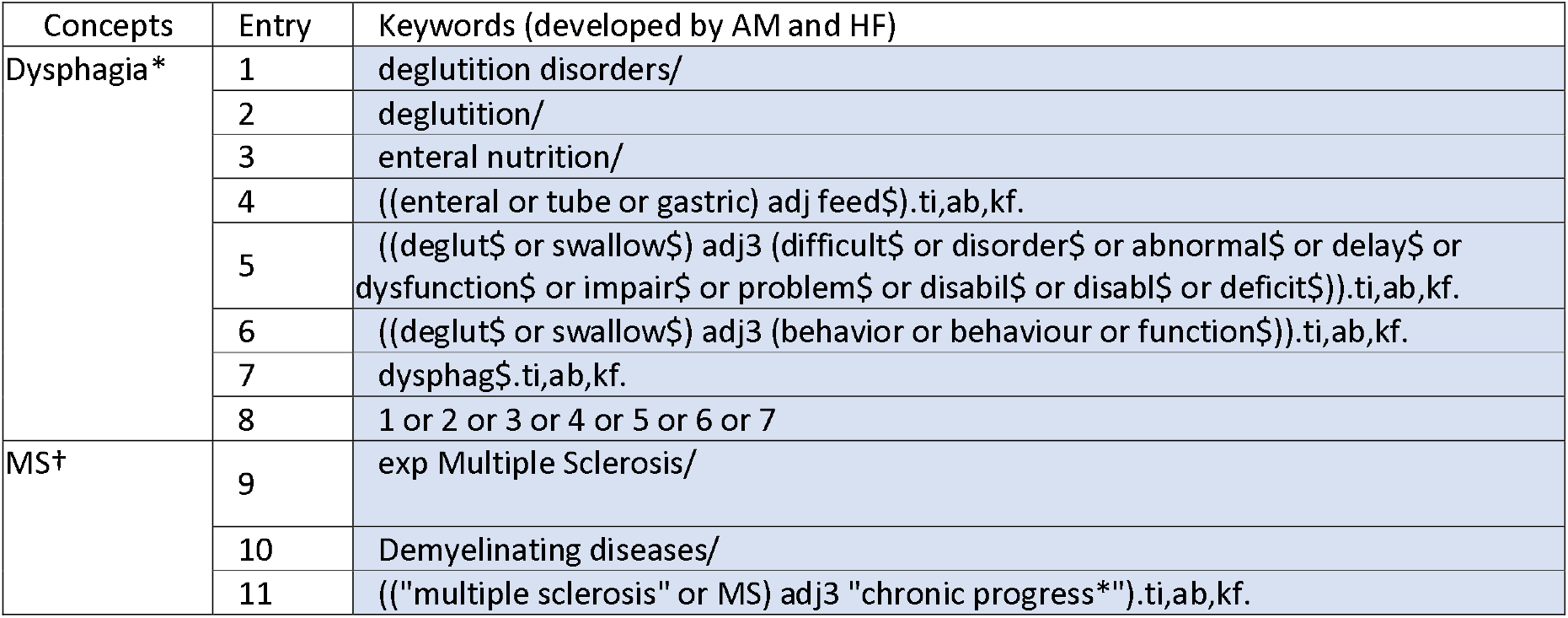
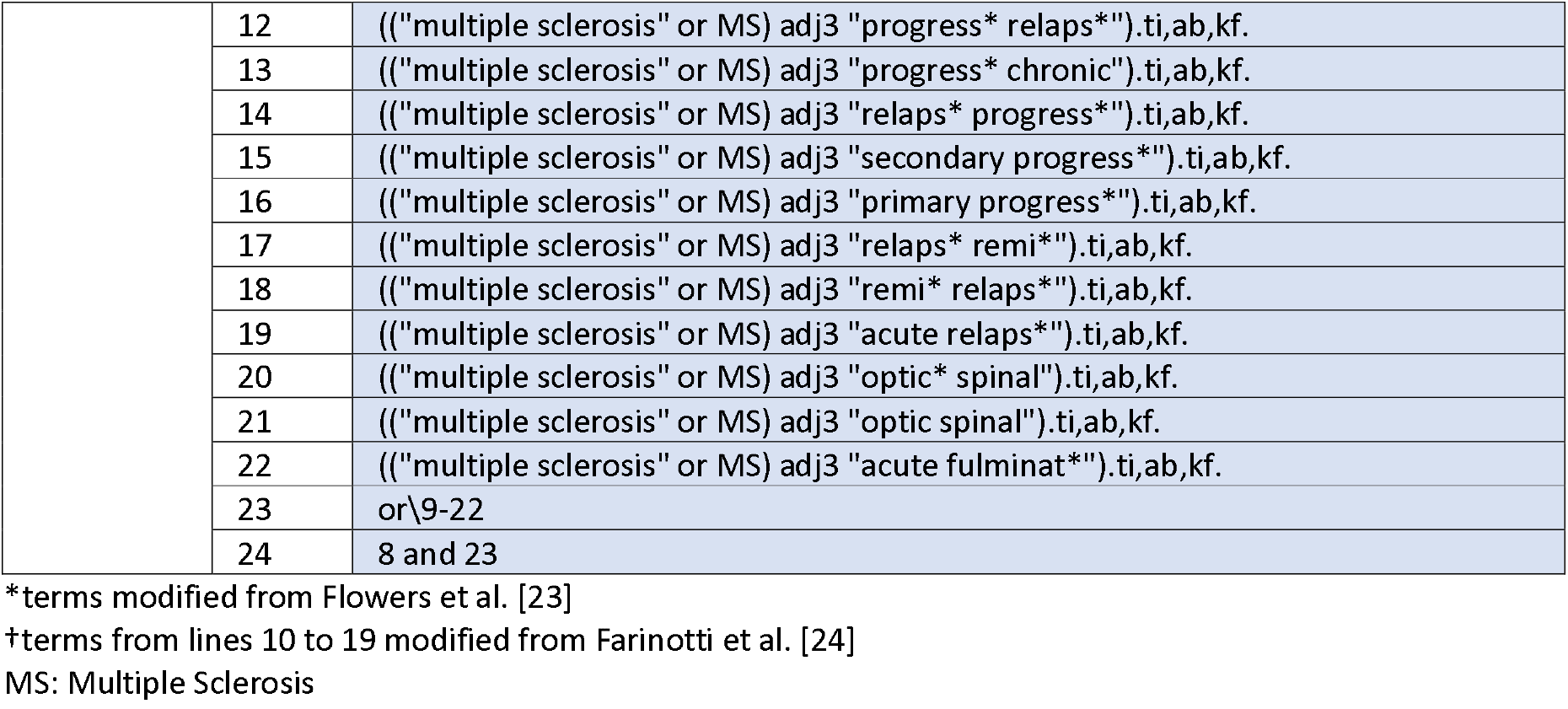
MEDLINE search terms for the concepts dysphagia and multiple sclerosis

### Eligibility criteria

Studies will be considered for inclusion if they have observational intent and involve retrospective or prospective consecutive or randomly selected sampling (either from a particular cohort or population). Study designs may include case series, cross-sectional, longitudinal, case–control, and/or other observational investigations as well as the control arm of randomized controlled trials. We will consider studies with at least 30 adults (18 years or older) with MS. Studies must include an aim to identify risk factors for oropharyngeal dysphagia (OPD). Corresponding studies that include follow-up time points will contribute to our interest in long-term outcomes.

During our review of abstracts and full articles, we will apply pre-defined exclusion criteria. That is, we will exclude studies involving convenience samples, those without extractable data for our outcomes of interest, and those reporting duplicate data. Any abstracts without corresponding full study publications will be excluded. We will also exclude articles without a clear sample of at least 30 pwMS and corresponding OPD (for at least a declared portion of the sample), identified by clinical or instrumental swallowing assessments. Finally, articles will be excluded if they do not conform to our operational definitions of OPD and MS or if they refer to oropharyngeal dysphagia induced by causes other than multiple sclerosis. We will contact authors when we cannot find full articles or when we wish to elucidate study characteristics such as method of dysphagia assessment. Our full systematic review reporting will conform to the Strengthening the Reporting of Observational Studies in Epidemiology (STROBE) checklist [25].

### Data collection

Study selection from primary articles will be performed in two stages:

I - Initial screening and coding of titles and abstracts whereby relevant abstracts (stage 1) will undergo full article review (stage 2) (Table 2);

**Table 2.**
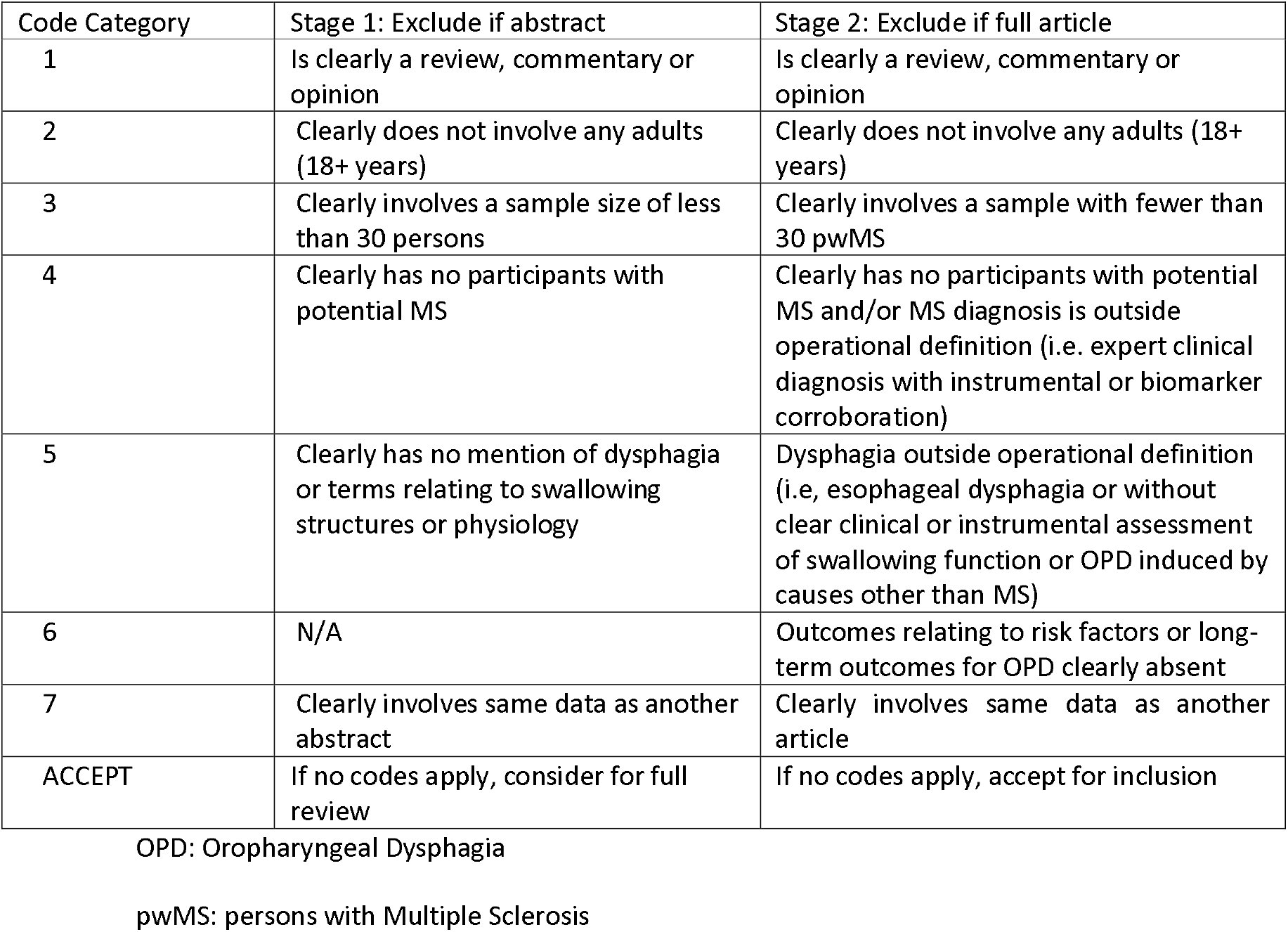
Proposed hierarchical coding categories for abstract and full article review

| Code Category | Stage 1: Exclude if abstract | Stage 2: Exclude if full article |
| --- | --- | --- |
| 1 | Is clearly a review, commentary or opinion | Is clearly a review, commentary or opinion |
| 2 | Clearly does not involve any adults (18+ years) | Clearly does not involve any adults (18+ years) |
| 3 | Clearly involves a sample size of less than 30 persons | Clearly involves a sample with fewer than 30 pwMS |
| 4 | Clearly has no participants with potential MS | Clearly has no participants with potential MS and/or MS diagnosis is outside operational definition (i.e. expert clinical diagnosis with instrumental or biomarker corroboration) |
| 5 | Clearly has no mention of dysphagia or terms relating to swallowing structures or physiology | Dysphagia outside operational definition (i.e. esophageal dysphagia or without clear clinical or instrumental assessment of swallowing function or OPD induced by causes other than MS) |
| 6 | N/A | Outcomes relating to risk factors or long-term outcomes for OPD clearly absent |
| 7 | Clearly involves same data as another abstract | Clearly involves same data as another article |
| ACCEPT | If no codes apply, consider for full review | If no codes apply, accept for inclusion |

OPD: Oropharyngeal Dysphagia
pwMS: persons with Multiple Sclerosis

II - Evaluation and coding of full articles for inclusion in the final sample (Table 2)

The review process will be conducted by two independent reviewers (blind to each other’s coding) across the two stages. Any discrepancies will be resolved by consensus discussion between the two reviewers and, when agreement is not possible, a third reviewer (also a member of the research team) will read the abstract or article independently and contribute to a decision. All references for the excluded articles will be retained for documentation purposes.

One data extractor will identify pertinent information from the final set of included articles and compile it into a table or spreadsheet. Extracted data will be verified by a second independent reviewer to ensure the accuracy of information from the following categories:

i. Study characteristics: first author’s name, year of publication, country in which the study was conducted, study design, and size of the sample.
ii. Study population and participant characteristics: age, gender, MS type, disease duration, and EDSS score.
iii. Diagnostic assessments for MS and dysphagia
iv. Risk factors for dysphagia: related to MS (e.g., MS type, disease duration, and EDSS score), patient characteristics (e.g., age, gender, and smoking or alcohol abuse), and/or comorbidities (e.g., psychological symptoms, cognitive impairments, or dysarthria)
v. Follow-up assessments of dysphagia
vi. Frequency and impact (e.g., severity) of detrimental medical (e.g., aspiration pneumonia, dehydration, malnutrition, institutionalization, and mortality), activity/participation (e.g., fewer social engagements around meals), or quality of life outcomes.

### Risk of bias in individual studies

We will apply appropriate risk of bias evaluations [26] such as the Newcastle-Ottawa Tool (NOS) [27] for observational studies (tool as our quality evaluation of included articles). To illustrate, the NOS contains grading for categories of selection (e.g., sample representativeness), comparability, (e.g., evaluation of confounders), and outcome (adequacy of follow-up period).

Further, if warranted, the Cochrane Collaboration’s Risk of Bias tool will be used for randomised controlled trials, based on the domains: sequence generation, blinding of participants, blinding of outcome measurement, allocation sequence concealment, missing data, selective outcome reporting, and other biases such as sources of funding and conflicts of interest [28]. For either type of quality appraisal (observational study quality scale or Cochrane’s risk of bias tool), two authors will independently review the included studies and resolve discrepancies by discussion and consensus agreement within the review team.

### Data analysis

We will provide a descriptive synthesis of the findings from the included studies, structured around target population characteristics, the type of assessments, and outcomes of interest. We will consider meta-analyses if there is an adequate number of studies and homogeneity of study populations and assessment methods. Otherwise, we will present a narrative synthesis of the results. We anticipate that there will be restricted scope for meta-analysis due to differing study populations and/or assessment methods along with a paucity of existing literature. Where studies have similar sample characteristics (including potential comparison groups), assessment methods, and corresponding outcomes, we will pool the results and apply various types of meta-analyses such as mean difference or standard mean differences for continuous outcomes and risk ratio measurement for categorical outcomes, with their 95% confidence intervals (CIs) depicted in forest plots.

#### Assessment of heterogeneity

If there is reason to consider meta-analysis, analyses will be performed using Cochrane’s Revman tool. Heterogeneity among primary studies will be assessed with the I^2^ statistic [28]. We will interpret the I^2^ statistic using the following guide: mild (between 0% and 24.9%), moderate (between 25% and 49.9%), severe (between 50% and 74.9%), and highly severe (between 75% and 100.0%). Nevertheless, we will subjectively evaluate possible sample heterogeneity before conducting a pooled analysis. Subsequently, if we proceed to apply the I2 statistic and heterogeneity greater than or equal to 25% is evident, we will present a qualitative synthesis of the findings.

#### Analysis of subgroups or subsets

If sufficient data are available, subgroup analyses may be conducted for different OPD assessment methods (e.g., clinical bedside evaluation, fiberoptic nasoendoscopic evaluation, or videofluoroscopic evaluation) relative to predictive factors and/or long-term outcomes.

#### Assessment of publication bias

Publication bias will be evaluated using a funnel plot (ie, plots of study results against precision) and Begg’s and Egger’s tests if an adequate number of studies (≥10) are identified.

## Discussion

Our search strategy is extensive compared to other recent systematic reviews in the field of multiple sclerosis [1,29] given the inclusion of numerous sources and comprehensive search terms. We believe that it will yield a broad capture of abstracts internationally but that many articles will derive from western or developed countries. This is important because many underrepresented countries, such as Iran, have a high and increasing prevalence of pwMS in certain regions [29,30].

Other systematic reviews have undertaken different lines of inquiry such as investigating the prevalence of dysphagia in pwMS without considering risk factors [1] or long-term outcomes [1,9]. Thus, we will extend the knowledge base in a new content area (involving predictors and long-term outcomes of dysphagia in pwMS). Thus, identification of literature in the field of MS will provide new insights into the repercussions of dysphagia and offer direction for the development of screening protocols and improved management in pwMS. It will anticipate that as the protocol consider multiple predictors (e.g., related to MS, patient characteristics, and/or comorbidities) and outcomes (e.g., medical, activity/participation, or quality of life), the results of our systematic review may be report in multiple publication paper.

We anticipate that various limitations will result during our search of the literature. First, studies may not report the timeframe between dysphagia onset, assessments, and associated outcomes. Second, dysphagia identification in studies might be based on cursory screenings, patient self-report nonstandardized questionnaires, and/or subjective clinical assessments rather than from instrumental reference tests such as videofuoroscopy and fiberoptic nasoendoscopy. Finally, it may be difficult to pool results from the existing literature for some of the predictors or outcomes if investigations restrict enrolment to particular types of pwMS, involving subsamples of larger studies, or fail to incorporate shared definitions and research methods in the field of multiple sclerosis [31].

## Conclusion

Although the frequency of dysphagia in pwMS has been considered within the past two decades [1,9], a poor understanding of associations to disease-related predictors and negative outcomes remains. Our proposed systematic review will address such a gap in the literature, as we will attempt to elucidate predictors of dysphagia and long-term outcomes from observational studies reporting frequencies of dysphagia over the long-term. Where relevant, we will pool results across studies or extract individual-level data that may permit us to model predictors of dysphagia and/or its associated long-term outcomes in pwMS. Our inquiry will offer the means to inform best practices in the early detection of dysphagia and provide information that can be incorporated into guidelines and clinical practice initiatives for the management of dysphagia in pwMS.

## Data Availability

All data produced in the present study are available upon reasonable request to the authors

## Acknowledgements

We acknowledge all institutions that funded our research.

## Data availability

All data produced in the present study are available upon reasonable request to the authors.

## References

[1] X.L. Guan, H. Wang, H.S. Huang, L. Meng, Prevalence of dysphagia in multiple sclerosis: a systematic review and meta-analysis, Neurological Sciences. 36 (2015) 671–681. https://doi.org/10.1007/S10072-015-2067-7/METRICS.

[2] C. Tassorelli, R. Bergamaschi, S. Buscone, M. Bartolo, A. Furnari, P. Crivelli, E. Alfonsi, E. Alberici, G. Bertino, G. Sandrini, G. Nappi, Dysphagia in multiple sclerosis: From pathogenesis to diagnosis, Neurological Sciences. 29 (2008) 360–363. https://doi.org/10.1007/S10072-008-1044-9/METRICS.

[3] R. Marchese-Ragona, D.A. Restivo, G. Marioni, G. Ottaviano, S. Masiero, A. Staffieri, Evaluation of swallowing disorders in multiple sclerosis, Neurological Sciences. 27 (2006) s335–s337. https://doi.org/10.1007/S10072-006-0654-3/METRICS.

[4] P. Calcagno, G. Ruoppolo, M.G. Grasso, M. De Vincentiis, S. Paolucci, Dysphagia in multiple sclerosis – prevalence and prognostic factors, Acta Neurol Scand. 105 (2002) 40–43. https://doi.org/10.1034/J.1600-0404.2002.10062.X.

[5] M. Prosiegel, A. Schelling, E. Wagner-Sonntag, Dysphagia and multiple sclerosis., Int MS J. 11 (2004) 22–31. https://europepmc.org/article/med/15125813 (accessed April 21, 2023).

[6] O. Ekberg, S. Hamdy, V. Woisard, A. Wuttge-Hannig, P. Ortega, Social and psychological burden of dysphagia: Its impact on diagnosis and treatment, Dysphagia. 17 (2002) 139–146. https://doi.org/10.1007/S00455-001-0113-5/METRICS.

[7] R.J.C.G. Verdonschot, L.W.J. Baijens, I. van de Kolk, B. Kremer, R.J.C.G. Verdonschot, L.W.J. Baijens, B. Kremer, S. Vanbelle, C. Leue, Affective symptoms in patients with oropharyngeal dysphagia: A systematic review,J Psychosom Res. 97 (2017) 102–110. https://doi.org/10.1016/J.JPSYCHORES.2017.04.006.

[8] A.D. Sadovnick, K. Eisen, G. Ebers, D.W. Paty, Cause of death in patients attending multiple sclerosis clinics, Neurology. 41 (1991) 1193–1193. https://doi.org/10.1212/WNL.41.8.1193.

[9] A. Aghaz, A. Alidad, E. Hemmati, H. Jadidi, L. Ghelichi, Prevalence of dysphagia in multiple sclerosis and its related factors: Systematic review and meta-analysis, Iran J Neurol. 17 (2018) 180. https://doi.org/10.18502/ijnl.v17i4.592.

[10] F.J. Thomas, C.M. Wiles, Dysphagia and nutritional status in multiple sclerosis, J Neurol. 246 (1999) 677–682. https://doi.org/10.1007/S004150050431/METRICS.

[11] L. Hartelins, P. Svensson, Speech and Swallowing Symptoms Associated with Parkinson’s Disease and Multiple Sclerosis: A Survey, Folia Phoniatrica et Logopaedica. 46 (1994) 9–17. https://doi.org/10.1159/000266286.

[12] A. De Pauw, E. Dejaeger, B. D’hooghe, H. Carton, Dysphagia in multiple sclerosis, Clin Neurol Neurosurg. 104 (2002) 345–351. https://doi.org/10.1016/S0303-8467(02)00053-7.

[13] W. Herrera, B.E. Zeligman, J. Gruber, M.C. Jones, R. Pautler, R. Wriston, M. Cain, T. Prescott, N. Cobble, J.S. Burks, Dysphagia in Multiple Sclerosis: Clinical and Videofluoroscopic Correlations, Neuro Rehab. 4 (1990) 1–8.

[14] Z. Sadeghi, M. Afshar, A. Ebadi, K. Baghban, Z.S. Qureshi, Swallowing Disorder in Multiple Sclerosis: Modified Version of the Screening Tool, Archives of Rehabilitation. 21 (2020) 236–255. https://doi.org/10.32598/RJ.21.2.3036.1.

[15] M. Tarameshlu, A.R. Azimi, L. Ghelichi, N.N. Ansari, Prevalence and predictors of dysphagia in Iranian patients with multiple sclerosis, Med J Islam Repub Iran. 31 (2017) 133. https://doi.org/10.14196/MJIRI.31.133.

[16] S.D. Pajouh, N. Moradi, M.J. Shaterzadeh Yazdi, S.M. Latifi, M. Mehravar, N. Majdinasab, A.R. Olapour, M. Soltani, F. Khanchezar, Diagnostic evaluation of dysphagia in multiple sclerosis patients using a Persian version of DYMUS questionnaire, Mult Scler Relat Disord. 17 (2017) 240–243. https://doi.org/10.1016/J.MSARD.2017.08.012.

[17] S. Abraham, L.C. Scheinberg, C.R. Smith, N.G. Larocca, Neurologic Impairment and Disability Status in Outpatients with Multiple Sclerosis Reporting Dysphagia Symptomatology,Neurorehabil Neural Repair. 11 (1997). https://doi.org/10.1177/154596839701100102.

[18] R. Bergamaschi, P. Crivelli, C. Rezzani, F. Patti, C. Solaro, P. Rossi, D. Restivo, D. Maimone, A. Romani, S. Bastianello, E. Tavazzi, E. D’Amico, C. Montomoli, V. Cosi, The DYMUS questionnaire for the assessment of dysphagia in multiple sclerosis, J Neurol Sci. 269 (2008) 49–53. https://doi.org/10.1016/J.JNS.2007.12.021.

[19] Z. Sadeghi, Z.S. Ghoreishi, H. Flowers, P. Mohammadkhani, F. Ashtari, M. Noroozi, Depression, Anxiety, and Stress Relative to Swallowing Impairment in Persons with Multiple Sclerosis, Dysphagia. 36 (2021) 902–909. https://doi.org/10.1007/S00455-020-10207-X/METRICS.

[20] B.J. Wang, F.L. Carter, K.W. Altman, Relationship between Dysarthria and Oral-Oropharyngeal Dysphagia: The present evidence, Ear Nose Throat J. (2020). https://doi.org/10.1177/0145561320951647/ASSET/IMAGES/10.1177_0145561320951647-IMG1.PNG.

[21] International Classification of Functioning, Disability and Health (ICF), (n.d.). https://www.who.int/standards/classifications/international-classification-of-functioning-disability-and-health (accessed April 21, 2023).

[22] C.M. Poser, D.W. Paty, L. Scheinberg, W.I. McDonald, F.A. Davis, G.C. Ebers, K.P. Johnson, W.A. Sibley, D.H. Silberberg, W.W. Tourtellotte, New diagnostic criteria for multiple sclerosis: guidelines for research protocols., Ann Neurol. 13 (1983) 227–231. https://doi.org/10.1002/ANA.410130302.

[23] H. Flowers, D. Bérubé, M. Ebrahimipour, M.F. Perrier, S. Moloci, S. Skoretz, Swallowing behaviours and feeding environment in relation to communication development from early infancy to 6 years of age: a scoping review protocol, BMJ Open. 9 (2019) e028850. https://doi.org/10.1136/BMJOPEN-2018-028850.

[24] M. Farinotti, L. Vacchi, S. Simi, C. Di Pietrantonj, L. Brait, G. Filippini, Dietary interventions for multiple sclerosis, Cochrane Database of Systematic Reviews. (2012). https://doi.org/10.1002/14651858.cd004192.pub3.

[25] E. von Elm, D.G. Altman, M. Egger, S.J. Pocock, P.C. Gøtzsche, J.P. Vandenbroucke, The Strengthening the Reporting of Observational Studies in Epidemiology (STROBE) Statement: Guidelines for reporting observational studies, International Journal of Surgery. 12 (2014) 1495–1499. https://doi.org/10.1016/J.IJSU.2014.07.013.

[26] L.L. Ma, Y.Y. Wang, Z.H. Yang, D. Huang, H. Weng, X.T. Zeng, Methodological quality (risk of bias) assessment tools for primary and secondary medical studies: what are they and which is better?, Military Medical Research 2020 7:1. 7 (2020) 1–11. https://doi.org/10.1186/S40779-020-00238-8.

[27] J. Peterson, V. Welch, M. Losos, P. Tugwell, The Newcastle-Ottawa scale (NOS) for assessing the quality of nonrandomised studies in meta-analyses, Ottawa: Ottawa Hospital Research Institute. 2 (2011) 1–12.

[28] J.P.T. Higgins, D.G. Altman, P.C. Gøtzsche, P. Jüni, D. Moher, A.D. Oxman, J. Savović, K.F. Schulz, L. Weeks, J.A.C. Sterne, The Cochrane Collaboration’s tool for assessing risk of bias in randomised trials, BMJ. 343 (2011). https://doi.org/10.1136/BMJ.D5928.

[29] O. Mirmosayyeb, N. Ebrahimi, A. Shekarian, A. Afshari-Safavi, V. Shaygannejad, M. Barzegar, S. Bagherieh, Prevalence of dysphagia in patients with multiple sclerosis: A systematic review and meta-analysis, Journal of Clinical Neuroscience. 108 (2023) 84–94. https://doi.org/10.1016/J.JOCN.2023.01.006.

[30] S. Bagherieh, O. Mirmosayyeb, S. Vaheb, A. Afshari-Safavi, M. Barzegar, F. Ashtari, V. Shaygannejad, Prevalence and Incidence of Multiple Sclerosis in Isfahan Province, Iran: A 25-Year Population-Based Study, Mult Scler Relat Disord. 71 (2023) 104310. https://doi.org/10.1016/J.MSARD.2022.104310.

[31] J.A. Cohen, M. Trojano, E.M. Mowry, B.M.J. Uitdehaag, S.C. Reingold, R.A. Marrie, Leveraging real-world data to investigate multiple sclerosis disease behavior, prognosis, and treatment., Mult Scler. 26 (2019) 23–37. https://doi.org/10.1177/1352458519892555.

